# Analysis and visualization of epidemics on the timescale of burden: derivation and application of Epidemic Resistance Lines (ERLs) to COVID-19 outbreaks in the US

**DOI:** 10.1101/2021.05.03.21256542

**Authors:** Alex Washburne, Justin Silverman, Jose Lourenco, Nathaniel Hupert

**Affiliations:** Selva Analytics LLC, Bozeman, MT, USA; College of Information Science and Technology, Pennsylvania State University, University Park, PA, USA; Department of Statistics, Pennsylvania State University, University Park, PA, USA; Department of Medicine, Pennsylvania State University, University Park, PA, USA; Institute for Computational and Data Science, Pennsylvania State University, University Park, PA, USA; Department of Zoology, University of Oxford, Oxford, UK; Weill Cornell Medicine and Cornell Institute for Disease and Disaster Preparedness, New York, NY, USA; The COVID-19 International Modeling (CoMo) Consortium, University of Oxford, Oxford UK

## Abstract

The 2020 COVID-19 pandemic produced thousands of well-quantified epidemics in counties, states, and countries around the world. Comparing the dynamics and outcomes of these nested epidemics could improve our understanding of the efficacy of non-pharmaceutical interventions (NPIs) and help managers with risk assessment across multiple geographic levels. However, cross-outbreak comparisons are challenging due to their variable dates of introduction of the SARS-CoV-2 virus, rates of transmission, case detection rates, and asynchronous and diverse management interventions.

Here, we present a graphical method for comparing ongoing COVID-19 epidemics by using disease burden as a natural timescale for comparison. Trajectories of growth rates of cases over the timescale of lagged deaths per-capita produces coherent visual comparisons of epidemics that are otherwise incoherent and asynchronous in the timescale of calendar dates or incomparable using non-stationary measures of burden such as cases. Applied to US COVID-19 outbreaks at the county and state level, this approach reveals lockdowns reducing transmission at fewer deaths per-capita early in the epidemic, reopenings causing resurgent summer epidemics, and peaks that while separated in time and place actually occur at points of similar per-capita deaths.

Our method uses early and minimally mitigated epidemics, like that in NYC in March-April 2020 and Sweden in later 2020, to define what we call “epidemic resistance lines” (ERLs) or hypothesized upper bounds of epidemic speed and burden. ERLs from less-mitigated epidemics allow benchmarking of resurgent summer epidemics in the US. In particular, the unmitigated NYC epidemic resistance line appears to bound the growth rates of 3,000 US counties and funnel growth rates across counties to their peaks where growth rates equal zero in the fall and winter of 2020. Corroboration of upper-bounds on epidemic trajectories allowed early predictions of mortality burden for unmitigated COVID-19 epidemics in these populations, predictions that were more accurate for counties in states without mask-wearing mandates. We discuss how this method could be used for future epidemics, including seasonal epidemics caused by influenza or ongoing epidemics caused by new SARS-CoV-2 variants.

**Press Summary:** Why, despite no statewide mask-wearing mandates or other restrictions like restaurant closures, did South Dakota’s COVID-19 epidemic peak not in January, when seasonal forcing wanes, but in early November? Why are we not seeing a resurgent epidemic in Florida or Texas, where non-pharmaceutical interventions have been relaxed for months? How can we compare the current outbreak in India with other countries’ epidemics to contextualize the speed of the Indian outbreak and estimate the potential loss of life?

We have developed a new method of visualizing epidemics in progress that can help to compare distinct COVID-19 outbreaks to understand, in specific cases like South Dakota, why they peaked when they did. The “when” in this case does not refer to prediction of a calendar date, but rather a point in the accumulation of deaths in a given locale due to the disease in question. The method presented in this paper therefore essentially uses population-based burden of disease as a timescale for measuring epidemics. Just as the age of a car can be measured in years or miles, the age of a COVID-19 epidemic can be measured in days or deaths per-capita. Plotting growth rates of cases as a function of per-capita deaths 11 days later produces a real-time visual comparison of epidemics that are otherwise asynchronous in time.

This approach permits both direct comparison across local outbreaks that may be disparate in time and/or place, as well as benchmarking of any outbreak against known exemplars of archetypal response strategies, such as New York City’s unmitigated urban outbreak in Spring 2020 and Sweden’s uncontained summer 2020 epidemic. Whether comparing the speed of resurgent outbreaks following relaxation in US states like Florida or the peak mortality burden in fall outbreaks across thousands of US counties with and without statewide mask-wearing mandates, this method offers a simple, intuitive tool for real-time monitoring and prediction capability connecting epidemic speed, burden, and management interventions. While our findings point to compelling epidemiological hypotheses for peaks in less-regulated states, future work is needed to confirm and extend our results predicting mortality burden at the peak of confirmed cases in the ongoing and evolving COVID-19 pandemic.

## Introduction

Throughout rushed and chaotic early response to an outbreak of a novel pathogen, typically marked by poor case ascertainment amidst a flurry of non-pharmaceutical interventions (NPIs), two questions dominate: Is it over? And, if not, how much worse can it get? Public health officials, medical care providers, and politicians alike seek answers to these questions to pace their response efforts and gauge when to release socially, economically, and even medically burdensome interventions such as “lockdowns” and other measures. Epidemics are often forecast with rolling regressions that ignore epidemiological principles such as immunity [1], compartment models reflecting epidemiological principles and capable of counterfactual analysis but simplifying population structure and relying on key parameters like the infection fatality rate or subclinical rates that may be non-identifiable under unknown case-ascertainment rates [2], or ensembles of many models that perform well over short-term forecasts but, due to their ensemble nature, lack mechanistic underpinnings and, due to their poor long-term performance, can’t reveal counterfactual scenarios such as relaxations of NPIs required to guide policy [3, 4]. Unfortunately, these standard approaches often fail to provide, in real-time and in a locally-adapted and easily comprehensible form, actionable information on which to base existentially important management decisions for thousands of managers overseeing outbreaks in their own jurisdiction. Here, we introduce a new analytical and data visualization approach to address this critical gap in pandemic emergency response.

SARS-CoV-2 emerged in China in late 2019 [5] and has since spread throughout the world, overwhelming health care systems and causing a significant loss of life. The US epidemic alone saw 355,000 fatalities from COVID-19 from February 2020 to January 1, 2021 [6]. After thousands of asynchronous local epidemics started across US counties, a dizzying flurry of NPIs were introduced at multiple societal levels, ranging from school closures to crowd size limits and mask-wearing mandates to full economic and social “lockdowns” such as shelter-in-place orders. Later, most US counties saw new cases begin to decline near the end of 2020 and early 2021 [6]. Some regions with few NPIs in place (e.g. like South Dakota or Florida) have seen cases decline at mortality burdens less than what many expected based on mitigation policies in place of containment policies, and estimated herd immunity thresholds and infection fatality rates. Currently, public health officials balancing the competing risks of COVID-19 disease burden against the costs of interventions wish to understand whether cases would continue to decline in their jurisdiction if costly NPIs are relaxed [7, 8]. As painfully illustrated by the current outbreak in India, where senior officials relaxed NPIs and cases declined for two months until an explosive resurgence of cases overwhelmed health care systems across India, the question of distinguishing between temporary lulls from natural endpoints is of critical importance, as are estimates of the potential for explosive growth of resurgent epidemics and the mortality burden of unmitigated outbreaks. Operationally, this requires a means of comparing local in-progress epidemics to archetypal (e.g. less-mitigated) epidemics and determining through a comparative epidemiological analysis whether mitigation efforts are necessary to prevent additional morbidity and mortality or are simply masking natural declines in outbreaks that have reached an endpoint corroborated by other, less-mitigated epidemics.

An analogy invoking the “potential epidemic energy” of a completely susceptible population can provide intuition for how to compare ongoing outbreaks. Imagine that the world’s epidemic-naïve population is sitting on the top of a hill. Local COVID-19 epidemics are analogous to individual carts rolling down a hill and the accumulation of infections is analogous to the declining altitude or potential energy of these carts. If blindfolded passengers were to independently ride 3,000 different carriages down a traumatically steep and bumpy hillside, with decelerations from each cart’s variably effective applications of brakes, then when the cart is finally slowing down the passengers may wonder: “Are we there yet?” If these passengers learned that the first carriages to charge down the hill without application of brakes arrived at a similar place despite having enough momentum to have carried them further, and if hundreds of later carriages with less reliance on brakes were also slowing down at a similar place, the wary passengers may be more inclined to believe the ride is over and relax accordingly. If, however, carts that had no brakes ran straight into a lake farther downhill, the continued application of brakes may be prudent. Many of the world’s public health officials are like drivers of these hypothetical carts, eager to assess if their local epidemics have reached a natural endpoint and determine the risks of relaxing the brakes of NPIs. New, easily interpretable means for real-time comparison of local epidemics to less-mitigated alternatives can assist in this real-time decision making.

One way for these officials to assess if an epidemic in their jurisdiction has reached a natural endpoint would be to assess whether their population’s exposure to the new pathogen is above an estimated herd immunity threshold (HIT). A crude way to do this is to estimate the basic reproductive number *R*_0_ of the pathogen from case growth rates and serial interval distributions [9] and compute the theoretical herd immunity threshold for a homogeneous population, *HIT* = 1 − 1/*R*_0_. Since many cases were not ascertained [10], one must sample the population for evidence of exposure and test whether the empirically obtained estimate of cumulative exposure is greater than the estimated HIT. If the cumulative incidence is greater than the HIT, then many would agree the epidemic’s recent decline would be due to herd immunity; if not, then this approach is inconclusive. For example, SARS-CoV-2 has an estimated R0 in the range of 2.5 to 5.7 [11, 12, 13] which imply a theoretical herd immunity threshold of 60-80% cumulative incidence under homogeneous populations. A serosurvey in New York City estimated a 23% cumulative incidence by March 29^th^ in one of the hardest-hit areas in 2020 [14], well below the estimated HIT, producing an inconclusive result that locked NYC into retaining costly interventions without disproving the hypothesis that NYC may have reached a natural endpoint defined by a lower seroprevalence than estimated under assumptions of homogeneous populations and high rates of seroconversion.

The inconclusive result of NYC illustrates a general point about the dubious management value of theoretical herd immunity thresholds for relaxation of NPIs in ongoing epidemics. Not only is it logistically difficult to obtain accurate real-time estimates of cumulative incidence in the setting of a dangerous outbreak, but also the simple model for calculating herd immunity threshold contains assumptions that may not hold in real populations. Heterogeneity in susceptibility and transmission potential at the individual level, which may prove difficult to quantify for local populations, can reduce the HIT below that of a homogeneous population used for simple HIT calculations [15, 16, 17, 18]. Furthermore, low case-detection rates [10, 19], false-negative serological assays due to a failure to seroconvert or loss of seropositivity [20, 21], which are increasingly evident in seronegative patients with T-cells recognizing SARS-CoV-2 [22, 23], can all result in cases and serosurveys underestimating cumulative incidence. Consequently, if populations are managed by the rule that NPIs should be relaxed if and only if the (logistically-taxing empirically estimated) cumulative incidence is above a theoretical homogeneous HIT, it’s possible that those populations will be perpetually locked into a state of vigilance for an epidemic whose real progress may already have produced sufficient immunity in combination with behavioral changes to mitigate future epidemic growth. In our cart analogy, if the bottom of the hill is 1,000m above sea level but passengers can only see their elevation and not their actual surroundings, then using a theoretical altitude of 0m above sea level as a prerequisite to release the brakes and open the doors makes no sense. The pertinent question is: how can public health and other officials learn, in real time, from neighboring regions’ outbreaks, the likelihood their epidemic reached a natural endpoint or that their cart could fall off another cliff without the further application of brakes?

In this paper, we present a method using disease burden as a timescale for comparing past and ongoing regional COVID epidemics, to estimate in real-time the potential speed of resurgent epidemics under relaxed NPIs, and, by quantifying the potential speed of resurgent epidemics as a declining function of disease burden, to identify burdens at which the potential speed is predicted to become zero. Using our approach, trajectories of well-documented, less-mitigated epidemics can serve as hypothesized upper-bounds of speed and burden such that resurgent epidemics following relaxation of interventions can be used to corroborate or reject the hypothesized bounds. Our method revolves around plotting COVID-19 case growth rates, *r*(*t*) – the most widely available and up-to-date estimate of transmission and the speed of ongoing COVID epidemics - as a function of lagged per-capita deaths, *D*(*t + τ*_*D*_) – the most widely available and comparable measures of cumulative exposures. Plotting the phase diagram of *r* as a function of lagged *D* yields visual comparisons of epidemics that reveal the efficacy of NPIs and the speed of resurgent epidemics.

Importantly, these visual outputs are easily interpretable by those unfamiliar with the inherent technical and mathematical details, allowing these graphical comparisons to be used as tools of communication to stakeholders, including the general public. We improve the interpretability of these plots by pointing out that per-capita deaths can serve as an alternative timescale for aligning epidemics that are otherwise asynchronous in calendar dates; much like the age of a car can be measured in years or miles, the age of an epidemic can be measured in days or cumulative burden of disease, such as attributable per capita mortality. Interpreting per-capita deaths as a timescale permits interpreting *r*(*D*) plots into trajectories of transmission over both time and cumulative incidence that can be read from left to right with rolling comparisons of local transmission to less-mitigated epidemics at earlier calendar dates when they were at a similar point of cumulative burden. We term the hypothesized upper bounds on *r*(*D*) plots “epidemic resistance lines” (ERLs) in connection to resistance lines from quantitative finance [24], and we present graphical and statistical methods to compare thousands of epidemics to these ERLs.

In this study, we apply the above approach to US COVID epidemics, setting the early and explosive NYC epidemic in March-April 2020 as an epidemic resistance line for US counties and other demographically similar populations. We show that, despite its relatively relaxed management strategy, Sweden’s summer epidemic was bounded above by NYC ERL, and that the NYC trajectory also bounded the case growth rates of summer epidemics across US states, including states with minimal interventions like Florida and South Dakota. Following its corroboration as an upper bound for the speed of resurgent US outbreaks in the summer, we hypothesized the NYC ERL would predict the endpoints of epidemics across US counties with few NPIs and subject to similar seasonal forcing of respiratory viral transmission as that encountered by NYC in March 2020.

Across fall outbreaks in US counties seeing a synchronized increase in case growth rates consistent with expectations from historical influenza-like illness data [25], the NYC ERL bounded the epidemic speeds of resurgent, seasonally-forced epidemics and predicted the mortality burden of the US epidemic as a whole at its seasonally forced winter peak. By predicting an upper quantile bound on growth rates and funneling them to a *D*-intercept where growth rates are equal to zero, the preceding, repeatedly corroborated NYC ERL predicted the central tendency of mortality burden corresponding to winter peaks across thousands of US counties, especially counties whose states (e.g. South Dakota) did not implement mask-wearing mandates, close schools, or close restaurants. We discuss some interpretations of these results, the limitations of our work, and point to future work that may improve the robustness and utility of this method for comparative epidemiology of novel pandemic pathogens.

## Results

### The Timescale of Burden in an Ongoing Pandemic

Any quantity changing monotonically over time – such as the cumulative ticks of a clock, the fraction of ^14^C isotopes in a biological sample, or the entropy of the universe – can serve as a way of measuring time. However, time as measured by dates is not always the most useful measure of time for a dynamical system under study. The age of a car, for example, can be measured in years but it’s often more practical to measure the age of a car in miles as the latter relates to the car’s experienced burden and risk of needing repair. Similarly, for ongoing epidemics, cumulative incidence and its corresponding disease burden may be a useful timescale. Disease burden in a pandemic increases monotonically with time and bounds transmission through population immunity, but cumulative incidence is often not known in real-time for technical or logistical reasons.

For many COVID-19 epidemics around the world, we don’t know the calendar date *t*_0_ when the epidemic started or the time interval, *t* − *t*_0_, that has elapsed since the start of the epidemic. However, across many regions and points in time, we have a reliable measure of deaths due to COVID-19. For example, we don’t know the time in the Italian epidemic when, on March 8, 2020, many northern provinces were locked down. Yet, we know that, by March 8, 366 Italians had died from COVID-19, or roughly 6 deaths per 100,000 capita. Belgium reached a similar point in the timescale of deaths by March 19, New York State by March 24^th^, Brazil by April 13^th^, India by June 11^th^, and so on. At different calendar dates, these epidemics were at a similar level of burden due to COVID-19. At different calendar dates, assuming a similar infection fatality rates across these regions, these epidemics were at a similar point along what could be considered a natural timescale of burden for analyzing and comparing epidemic properties such as transmission rate (Figure 1).

**Figure 1:**
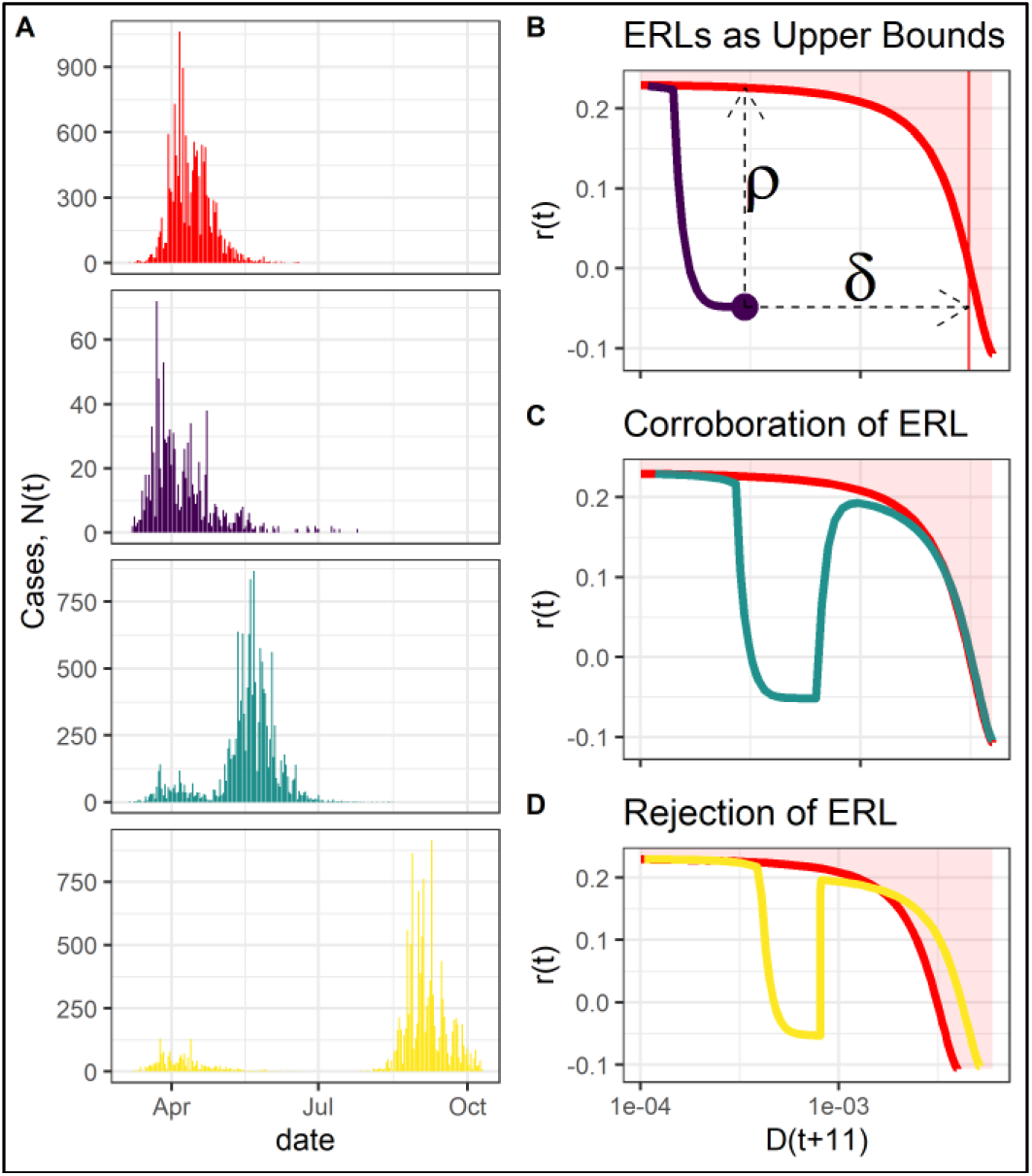
Epidemics on the timescale of deaths. **(A)** Case trajectories are often plotted over calendar dates. Plotted are four simulated epidemic scenarios: an unmitigated epidemic, a successfully contained epidemic, an epidemic that was contained but became unmitigated following relaxation of interventions, and an epidemic with a higher infection fatality rate than the other three. **(B)** Growth rates as a function of lagged deaths per-capita allow real-time graphical comparison of epidemics; these trajectories can be read from left to right as one reads epidemics over time. A successfully contained epidemic (purple line) can be compared to an unmitigated epidemic to provide estimates of resurgence potential speed *ρ* and mortalitiy, *δ*. **(C)** Unmitigated epidemics define “epidemic resistance lines”; here, the relaxation of a simulated epidemic corroborates the ERL. **(D)** An early, unmitigated epidemic & hypothesized ERL can be rejected by later epidemics that significantly exceed the ERL’s *r*(*D*) trajectory, crossing into a rejection region (shaded red area).

Formally, cumulative deaths or other measures of disease burden can be defined as a timescale for analysis by noting that cumulative deaths from COVID-19, *D*(*t*), increase monotonically with time. By imposing continuity in deaths through interpolation, any function of time during a COVID-19 epidemic can be converted into a function of deaths. In most epidemiological dynamical systems, finding exact functional form of *D*(*t*) or its inverse *t*(*D*) is difficult if not impossible with the contemporary mathematical tools. However, rather than working explicitly with *D*(*t*), it often suffices to work implicitly through its time derivative, *dD*/*dt*. For a simple model of an epidemic in a population compartmentalized into Susceptible *S*, Infected *I*, and Recovered/Resistant *R* populations, the dynamics of the infected population are given by

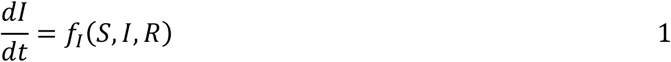

where *f*_*I*_ is some equation defining the rates of infection, recovery, and death. To utilize the timescale of deaths for analysis, we note that for a smooth function *D*(*t*), *dt*/*dD* = (*dD*/*dt*)^−1^, allowing us to use the chain rule to express the dynamic prevalence *I*(*D*) on the timescale of deaths as

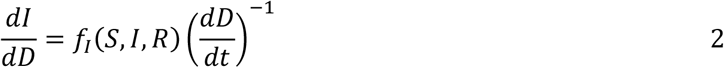

Since deaths increase nonlinearly over time, the timescale of deaths will warp our view of time of an epidemic, “slowing down” the epidemic where deaths are rapidly increasing (*dD*/*dt* large, its inverse small) and “speeding-up” time when the epidemic is otherwise slow-changing.

The timescale of deaths is a natural way to view and compare epidemics when there exists a nice relationship between deaths and cumulative incidence. A robust relationship between deaths and cumulative incidence can exist under a constant infection fatality rate or, where there is regional or temporal variation in IFR, if the relative infection fatality rates across regions and times can be estimated to produce risk-adjusted estimates of cumulative burden. If confirmed cases are detected at a rate proportional to disease prevalence, then the growth rate in cases, *r*(*t*), will decrease monotonically with cumulative incidence, *Z*(*t*). For an SIR model with density dependent transmission, *f*_*I*_ = (*βS* − *μ* − *γ*)*I* and

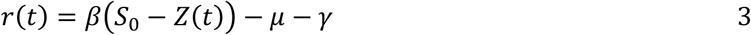

where *β* is the transmission rate, *S*_0_ the population size, *μ* the mortality rate of infected individuals and *γ*^−1^ the infectious period. For many simple epidemiological models, lagged deaths, *D*(*t + ι*), are approximately proportional to current cumulative incidence *Z*(*t*) (see SI: “Analysis of epidemics on timescale of deaths”). One can express the expected growth rate in cases for the SIR model above as

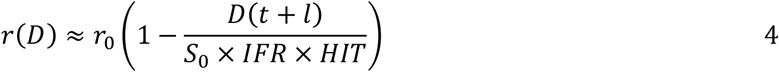

where *ι* = (*γ + μ*)^−1^ is the average time a person is infected and connects deaths today as a measure of cumulative incidence at the time of exposure, *IFR* is the infection fatality rate and *HIT* is the herd immunity threshold i.e. the attack rate at the peak of an unmitigated epidemic. Appropriately lagged deaths serve as a timescale such that when *D*(*t + ι*) = 0 the epidemic is at its maximum rate of growth and when the population fatality rate *D*(*t + ι*)/*S*_0_ = *IFR* × *HIT*, the epidemic is at its peak regardless its precise start-date.

Growth rates over the timescale of deaths defined in equation 4, what we call *r*(*D*) trajectories, define a curve from fast growth at zero burden to zero growth at the peak of an unmitigated epidemic. While herd immunity thresholds as extracted from epidemiological models is defined at a single point at which growth rates equal zero, this curve for unmitigated epidemics traces, among other things, an empirical estimate of the contribution of population immunity to reduced transmission throughout the entire epidemic without making strong assumptions about population structure, modes of transmission, and more. If NPIs and behavioral changes cause transmission rates, *β*(*t*), to decrease erratically from its maximal, unmitigated value, the *r*(*D*) trajectory for a mitigated epidemic will remain underneath that of an unmitigated epidemic. We thus call *r*(*D*) trajectories for minimally mitigated epidemics “epidemic resistance lines” (ERL) in reference to ‘resistance lines’ of quantitative finance [24]. For an epidemic halted via NPIs at a death burden, *D*_*c*_, one can use epidemic resistance lines, *r*_*ERL*_(*D*), from less-mitigated epidemics to estimate the maximum speed of a resurgent epidemic, *ρ* = *r*_*ERL*_(*D*_*c*_) and lives saved (or burden remaining) up to that point in the outbreak *δ* = *D*_*c*_ − *D*^*\**^ where *D*^*\**^ defines the burden at peak that satisfies *r*_*ERL*_ (*D*^*\**^) = 0 (Figure 1B). ERLs can be corroborated by resurgent epidemics tracing the *r*_*ERL*_(*D*) trajectory (Figure 2C) and rejected with the accumulation of epidemics that on average exceed the epidemic resistance line (Figure 2D). For more information, see: SI “Analysis of epidemics on the timescale of deaths”.

**Figure 2:**
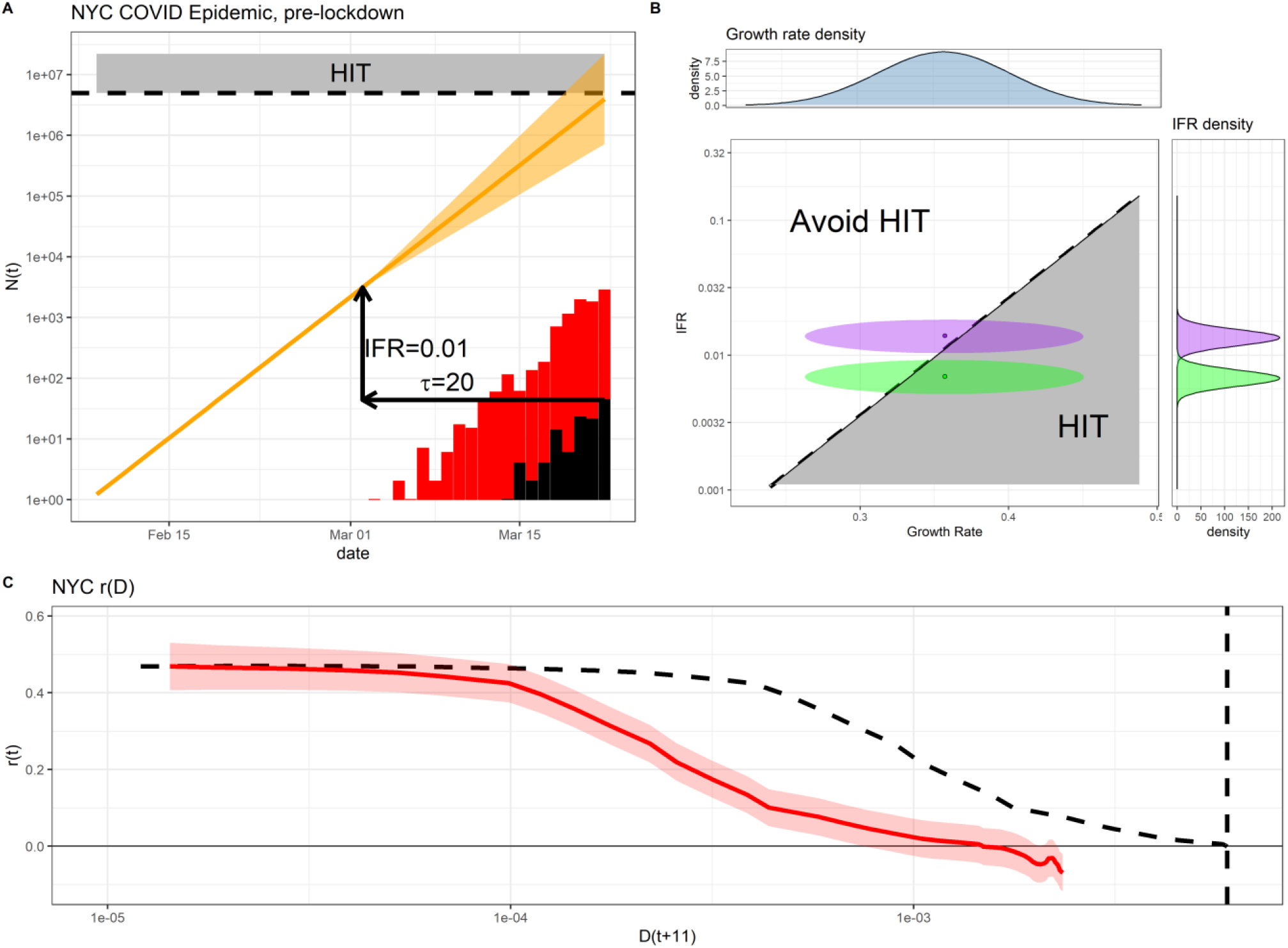
The NYC Epidemic as an ERL. **(A)** The NYC epidemic exhibited rapid and sustained exponential growth in cases (red) and deaths (black) up to the March 22 lockdown. A *τ* = 20 day lag from exposure to death, a 1% IFR, and estimated growth rates put 60% cumulative exposure by March 22 within reach. **(B)** Uncertainty in estimates of growth rates and IFR produce uncertainty in cumulative incidence by the day of the lockdown. Serosurveys may overestimate IFRs (purple) due to emerging evidence of seronegative patients with T-cells recognizing SARS-CoV-2. Adjusting for findings from T-cell surveys, lower IFRs (green) increase the probability of NYC reaching a 60% HIT. **(C)** We hypothesized the NYC epidemic could serve as an epidemic resistance line (red line) for later epidemics. A counter-factual ending at a 0.36% population fatality rate (black dashed line) corresponding to 60% attack rate and 0.6% IFR is included for comparison.

ERLs can serve as either strict upper bounds for strictly decreasing transmission, quantile bounds for fluctuating transmission that averages less than the unmitigated scenario, or expected trajectories for transmission and infection fatality rates fluctuating about the means of the unmitigated scenario. The existence of ERLs in plots of *r* versus *D* motivates the real-time comparison of ongoing epidemics’ *r*(*D*) trajectories with less-mitigated scenarios, and underlies the statistical analysis, visualizations, and interpretations in this paper. Early epidemics hypothesized to be unmitigated define a continuous *r*(*D*) curve predicting the maximum speed of resurgence following relaxation of NPIs and hypothesized endpoints for later unmitigated epidemics. The choice of which epidemics to compare requires careful reasoning based on a knowledge of pathogen transmission, local population structure (including risk factors that may affect the accumulation of disease burden), behavioral factors, seasonality, and interventions in place. Below, we motivate the use of NYC as an epidemic for comparison of later epidemics across US states, counties, and countries like Sweden with comparable demographics.

### The NYC Epidemic During March-April 2020

The first detected case of COVID in NYC was reported on February 29, 2020, after which exponentially growing new confirmed cases, ICU arrivals across providers, and new deaths doubled roughly every 2 days (Figure 2A). By the time the New York lockdown came into effect on March 22, NYC reported 119 casualties, 44 of which occurred on the day the lockdown began. At the time of the lockdown, new cases and deaths were still growing exponentially (FIGURE 2A).

A simple exponential projection of an estimated exposed population reveals the plausibility of NYC hitting a postulated 60% HIT by March 22. Given lag *τ* = 20 days from exposure to death and a 1% IFR [26], the distribution of likely exponential growth rates produces a non-negligible chance that NYC could have reached this postulated HIT (Figure 2A). Projecting the growth of the exposed pool, *E*(*t*), from *τ* days prior to the date of the lockdown, *t*_*LD*_, under continued exponential growth can be estimated as

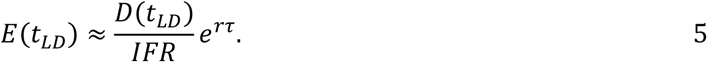

This equation is intuited in two steps: (1) (*D*_*LD*_/*IFR*) estimates the size of an exposed cohort by scaling up new deaths with the infection fatality rate, and (2) *e*^*rτ*^ project forward for *τ* days at growth rate, *r* (Figure 2A). If we set *E*(*t*_*LD*_) = *HIT* (here, 60% the population size of NYC), then we can solve for the IFR as a function of the growth rate, noting that exponential projections hit the herd immunity threshold if

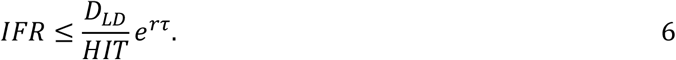

One study [27] estimated an IFR of 1.39% (95% CI 1.04-1.77%) for the NYC epidemic and serosurveys later estimating a 0.97% IFR for NYC [26] may have underestimate cumulative incidence and, consequently, overestimate the IFR. This is most evident in a study in the UK where for every seropositive patient, there was an additional seronegative patient with T-cells recognizing SARS-CoV-2 specific epitopes, indicating past infection and current cellular immunity [23]. Accounting for a 50% false-negative rate of serosurveys would approximately halve the estimated infection fatality rate compared to that estimate by raw seroprevalence estimates. If the model-based estimates over-estimate the IFR by a factor of two and the IFR were 0.7% (95% CI 0.52-0.88%), then 79% of these exponential projections of the NYC exposed population exceed the HIT. If the serosurvey over-estimated IFR by a factor of two, then 87% of these projections exceed the HIT.

These projections have many limitations, including potential biases in death-ascertainments in the early pandemic, the over-simplicity of exponential projections, the dependence of HIT on *r* is not accounted for, and more. Our goal with these simple projections is not to lean heavily on the resultant probabilities, but rather to make the argument that it’s plausible the NYC COVID-19 epidemic could have reached a natural endpoint for SARS-CoV-2 variants circulating at the time, and that the associated probability ultimately depends on several parameters: e.g. the attack rate at this “natural endpoint”, the growth rate, infection fatality rate, the lag from onset to death for these early infections, and any common non-stationarity in the ascertainment rate of cases and deaths in the early epidemic of NYC (Figure 2B).

Based on the plausibility of NYC hitting this early herd immunity threshold, we used the *r*_*NYC*_ (*D*) trajectory from NYC as a hypothesized ERL (Figure 2C) and monitored whether the NYC ERL defined upper quantiles of the speed of resurgent outbreaks and the average population fatality rates at the endpoints of later, less-mitigated epidemics subjected to similar seasonal forcing of transmission as NYC encountered in March. Using a negative binomial state-space model to estimate the growth rates of cases over time, we estimate the NYC epidemic peaked on April 7^th^ with an 11-day lagged deaths per-capita following peak cases,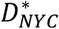, of approximately 1.5 deaths per 1,000 capita.

### Summer epidemic speed and endpoints following relaxation of NPIs

In March-April 2020, shelter-in-place orders were implemented across US states (SI: intervention timelines in focal US states). Many states relaxed their interventions early in the summer and experienced subsequent summer epidemics. While many countries and states attempted to contain their COVID-19 epidemics, Sweden embraced a mitigation strategy focused on reducing the exposure risk of elderly populations and encouraging voluntary social distancing [28] but otherwise imposed no mask-wearing mandates or shelter-in-place orders. By June 17^th^, we estimate cases in Sweden peaked near 1 death per 2,000 capita, well under 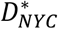. The early, less-mitigated epidemics in NYC and Sweden provide useful trajectories for comparison and understanding US states’ trajectories up to September 1, especially when including information on each state’s trajectory by interventions at the time [29].

US states and Sweden saw significant reductions in transmission early in their epidemics that reduced their case growth rate well under *r*_*NYC*_ (*D*) (Figure 3). After reopening in the summer, case growth rates rose across US states and all states had maximum resurgent epidemic speeds, *ρ*, bounded above by the NYC ERL (Figure 2). Florida and Georgia never reversed their relaxation of NPIs in the summer, yet their growth rate trajectories remained bounded by the New York line. These less-mitigated summer epidemics peaked later, in July and August, at a similar summer mortality burden of Sweden’s mid-June peak. Louisiana exceeded both the Swedish and the New York State trajectories but bounced off *r*_*NYC*_ (*D*) and reached its summer peak without any subsequent interventions, further corroborating the use of the NYC ERL.

**Figure 3:**
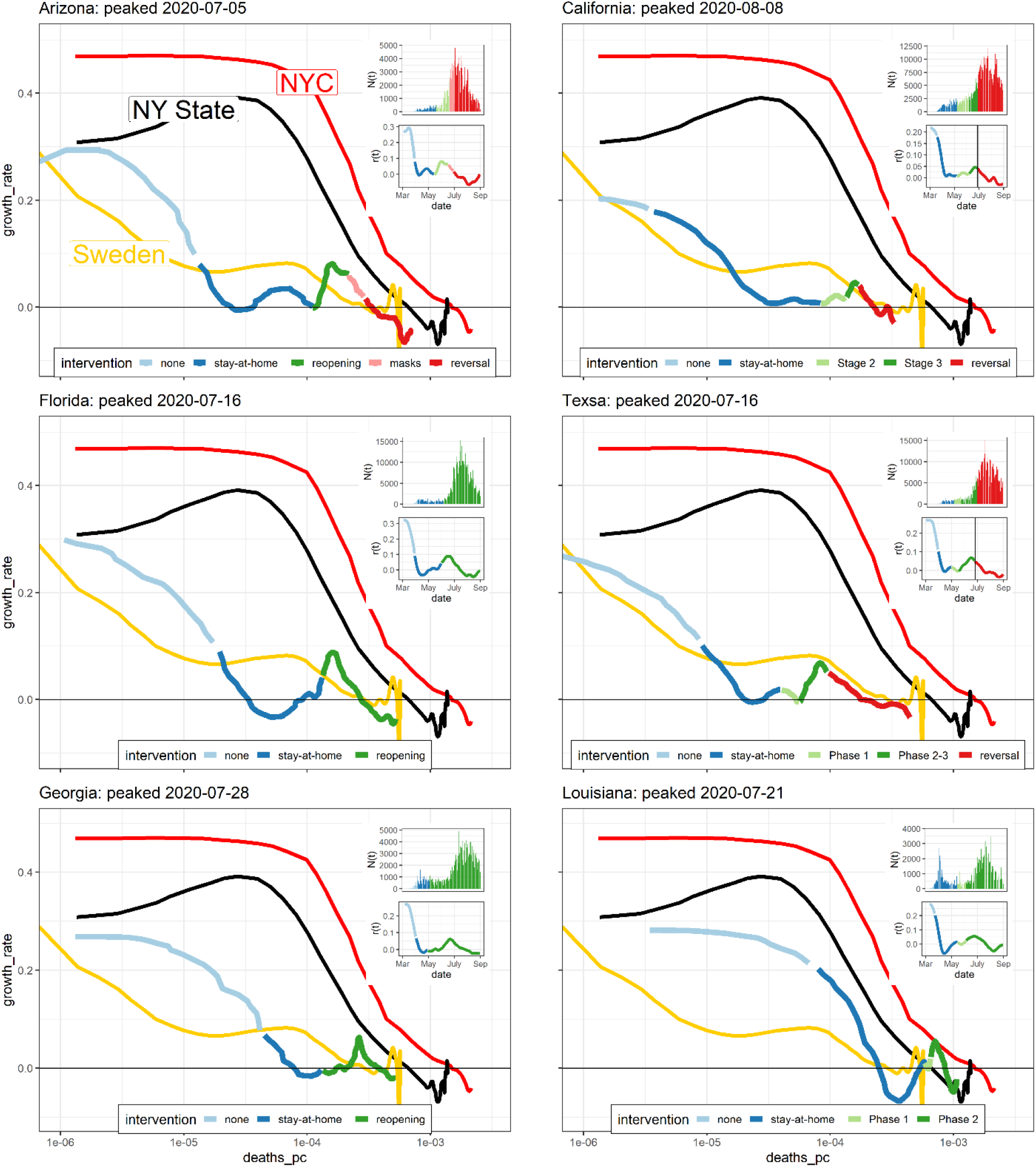
Epidemic resistance lines of summer peaks. *r*(*D*) plots for 6 US states with large summer epidemics compared to earlier-peaking trajectories from Sweden, New York State, and New York City. The focal state’s trajectory is colored by the contemporary intervention in that state. Inset plots show case counts and growth rates as a function of calendar dates for comparison. Behavioral changes and successful interventions produced lower growth rates than earlier New York state or NYC epidemics. Following reopening, growth rates of cases rose and deaths accumulated, but growth rates as a function of deaths per-capita were bounded by the NY State line and converged to peaks near Sweden’s less-mitigated Summer epidemic endpoint. In this way, the trajectory of the earlier and less-mitigated Swedish epidemic foreshadowed the mortality burden of local endpoints of later, less-mitigated US summer epidemics. Louisiana is an important exception - its excess of the NY State line was followed by a later peak at almost the exact point when Louisiana’s largest county’s trajectory bounced off the NYC line.

By mid-August, most US states had peaked, with peaks converging around 1 death per 2,000 capita consistent with the less-mitigated Swedish epidemic but significantly less than the 1.5 deaths per 1,000 capita predicted by the NYC line. The corroboration of the NYC epidemic as an upper bound for US state-level epidemics in the summer, most notably Louisiana which exceeded the Swedish and New York state lines, motivated the continued use of the NYC bound as a hypothesized ERL for the fall.

### US Counties’ Fall epidemics in relation to the NYC ERL

Starting in mid-August, a clear and synchronized increase in the growth rate of cases occurred across all US counties at almost the exact time that an increase in influenza-like illness visits to outpatient providers based on prior years’ records (Figure S1). During this period of probable seasonal forcing of transmission, many states remained open with relatively relaxed interventions, allowing us to test the appropriateness of the NYC ERL as a boundary for these seasonally-forced outbreaks across over 3,000 US counties. If the NYC epidemic defines an ERL, then it should be an upper-bound for epidemic speed in counties with transmission-reducing interventions in place. If NYC is bounds epidemic speeds, it should also bound later, epidemics’ steady decelerations to peaks where 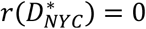, thereby predicting the per-capita deaths at peak, *D*^*\**^, for later unmitigated epidemics. Throughout, we refer to “peaks” as peaks in confirmed cases where *r*_*i*_(*t*^*\**^) = 0 for some county *i* where case-loads at the peak are at least three times the case loads of the preceding valley; “deaths per-capita at peak” refers to the corresponding lagged per-capita deaths *D*^*\**^ = *D*(*t*^*\**^ *+* 11) for a peak identified on day *t*^*\**^.

Cook, Harris, Los Angeles, Maricopa, and Miami-Dade counties – large urban centers and the five most populous US counties outside of NYC – all experienced asynchronous fall outbreaks whose *r*(*D*) trajectories mean-reverted along the NYC ERL, corroborating the NYC ERL. Cook County experienced its most prominent peak on November 11^th^ with 1.24 deaths per 1,000 capita, providing early additional corroboration and evidence the NYC ERL and the 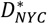 estimate of 1.5 deaths per 1,000 capita at peak may hold for the remainder of US counties peaking later during these seasonally-forced epidemics (Figure 4B). The deaths per-capita at later peaks of these large urban epidemics asymptotically approach the mortality burden predicted by the NYC epidemic resistance line (Figure 4C), and the maximum deaths per-capita at the most prominent peaks for these large urban counties was witnessed in Miami-Dade’s December 5^th^ peak occurring at 1.48 deaths per 1,000 capita, right under the hypothesized NYC upper bound.

**Figure 4:**
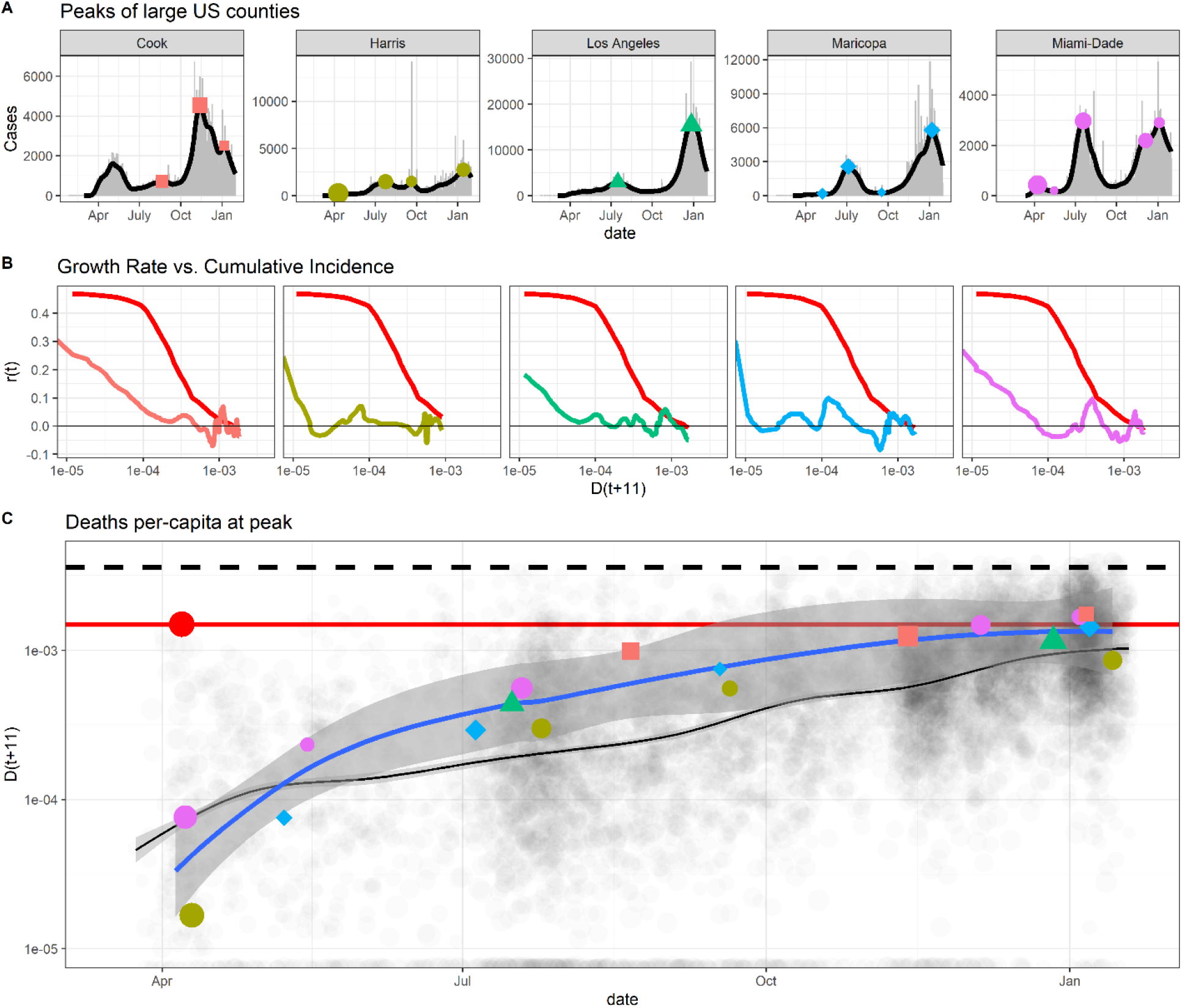
Deaths per-capita at peaks. **(A)** The top 5 most populous US counties, excluding NYC, had epidemics with multiple peaks, including fall epidemics that all corroborated the NYC line as an ERL. Peaks in the smoothed case trajectories are extracted and sized by prominence, measured as the ratio of cases at the peak to cases in the preceding valley. Points are colored & shaped uniquely for each county throughout this figure. **(B)**: These top-5 most populous counties all experienced outbreaks with early reductions in growth rates leading to large excursions below the NYC ERL, followed by seasonally-forced epidemics that rose to and descended to their peaks along the NYC ERL established prior by April 7, 2020. **(C)**: Deaths per-capita at multiple peaks in these top-5 most populous counties converge asymptotically over time to the endpoint defined by the NYC ERL. The blue line is the LOESS estimate for the five most populous counties' deaths per-capita at their multiple peaks; transparent black dots are all other US counties’ deaths per-capita at peaks and the black line is their LOESS estimate.

Across all US counties’ epidemics, the deaths per-capita at peak asymptotically approached approximately 1 death per 1,000 capita, confirming 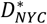 as an overestimate of mortality burden at the peaks of thousands of US county epidemics having some transmission-reducing policies & behaviors in place during their seasonally forced COVID epidemic (Figures 5A). During the 2020/2021 fall/winter COVID epidemic, the US had little in the way of national mandates and instead had a patchwork of state and county-level NPIs allowing the study the impact of NPIs on epidemic trajectories and mortality outcomes. Some states like South Dakota and Florida refrained from mask-wearing mandates and many other NPIs. Rather than attempt to control for the many confounding NPIs, we use statewide mask-wearing mandates as proxies for management intensity and attitudes towards social distancing across US counties. Plotting the COVID-19 deaths per-capita at peaks for states with and without state-wide mask-wearing mandates reveals the deaths per-capita at peak for states like South Dakota were higher than states with mask-wearing mandates (Figure 5B). Nonetheless, even these states had fewer deaths per-capita at their final peaks than predicted by the *r*_*NYC*_ (*D*) epidemic resistance line.

**Figure 5:**
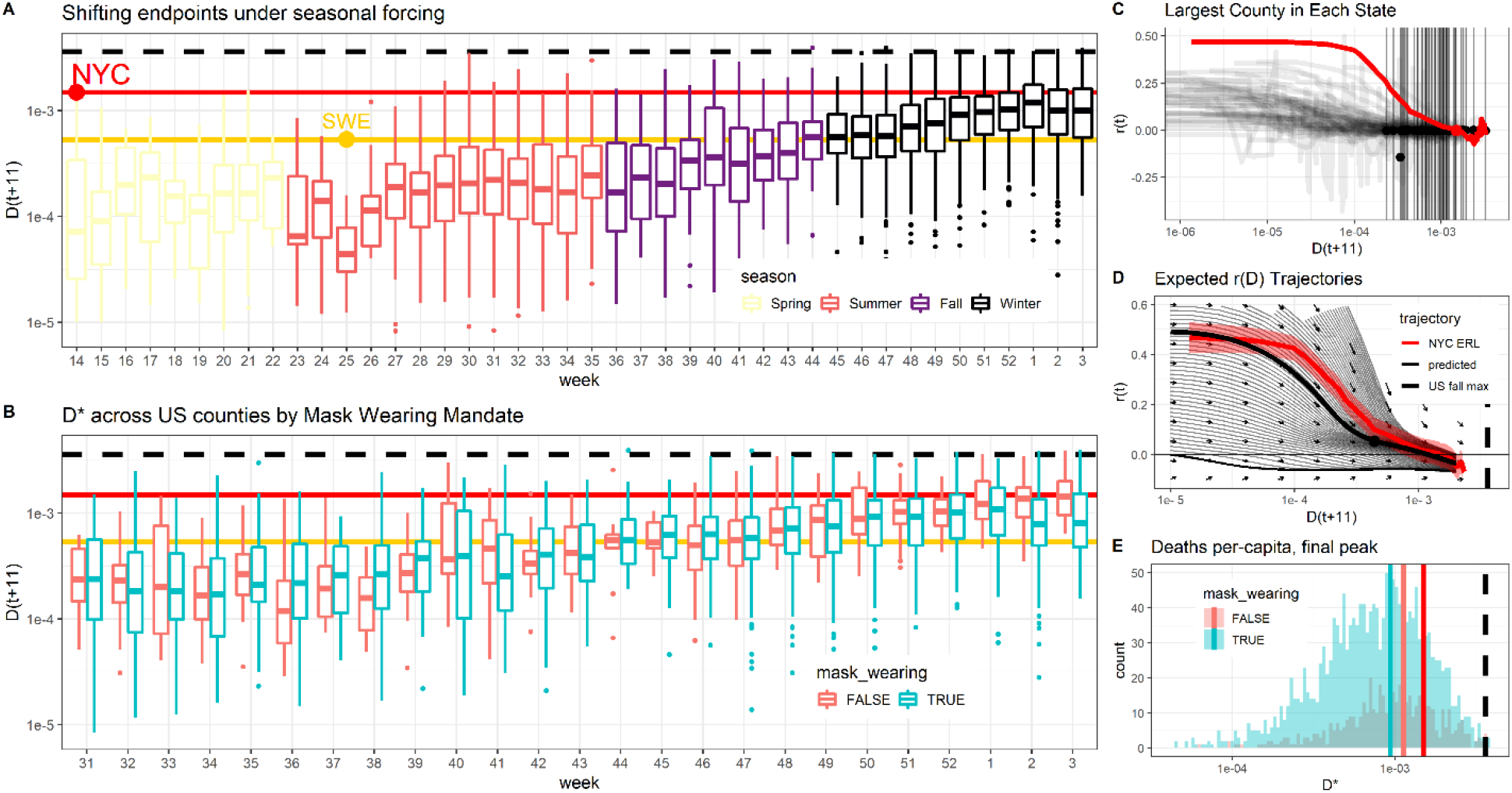
Global analysis of US counties’ epidemics in relation to the NYC ERL. **(A)** US county deaths per-capita 11 days after peak cases by week, colored by season. The stability of the distribution of *D*(*t +* 11) after peaks in cases underlies our predictions of summer peaks with the earlier and less-mitigated Swedish epidemic until week 40. Starting week 50 (December 8, 2020), the distribution of deaths per-capita at peak converges across weeks, with lower log-scale variance than in the summer, suggests this final point of arrival of US counties is an endpoint for the seasonally-forced COVID epidemic in the US. The upper quartile of boxplots for these final peaks falling near the NYC line is consistent with the NYC line being an ERL. **(B)** Using statewide mask-wearing mandates as a proxy for regulation intensity, we find less-regulated states converge to a distribution of *D*^*\**^ by December 2020 with interquartile ranges that include NYC predictions. The median deaths per-capita at peak in less-regulated states converges to 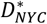 by January, providing suggestive evidence that the NYC line defines an estimate of the average death burden for a relatively unmitigated COVID epidemic. **(C)** The r(D) plots for the largest county in each state show a clear and nearly universal r(D) bound by the NYC line, and their final endpoints (vertical lines) fall in a distribution consistent with NYC being an upper bound for measures of central tendency (mean/median/mode) of deaths per-capita at peak in populous counties. **(D)** An inferred vector field of *r*(*D*) trajectories corroborates the NYC epidemic resistance line and numerical integration shows predicted trajectories funneled to peaks consistent with the NYC predictions. The predicted trajectory passing through the median maximum growth rate (and associated average deaths per-capita) of fall county epidemics falls under the NYC line. **(E)** Counties in states with statewide mask-wearing mandates had 14% reductions in mortality at peak relative to counties without, and we estimate *δ* = 29,000 US lives were saved in states with mask-wearing mandates.

The bulk of the distribution of *r*(*D*) trajectories among epidemics in the most populous counties across US states is contained underneath *r*_*NYC*_ (*D*) and their final peaks mean and median *D*^*\**^ fall under than 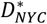 (Figure 5C), corroborating the epidemic resistance line of NYC as a quantile bound for these trajectories. A vector field of *r*′(*D*) trajectories inferred by a tensor spline reveals the average trajectories of all US counties in (*r, D*) space. Simulated trajectories on this vector field funnel towards peaks within the 95% credible regions of *r*_*NYC*_ (*D*), suggesting the average US county obeyed a trapping region defined by *r*_*NYC*_ (*D*). We estimate the maximum growth rate across all US counties, obtain the corresponding deaths per-capita for each county, and take the median of these fall maxima (*r, D*) values; the trajectory in our inferred *r*^′^(*D*) vector field passing through this midpoint of US fall maximum growth rates remains underneath *r*_*NYC*_ (*D*) (Figure 5D).

## Discussion

Recalling the analogy of carts losing potential epidemic energy while descending a hill, as the individual carts slow down and come to a rest on a flat plain, a detailed comparison of their trajectories with those of earlier, faster, and less mitigated carts can help passengers see whether or not they have indeed ended at a hypothesized bottom of the hill. By analyzing thousands of COVID-19 epidemics across US states and US counties, we found that the early epidemic of NYC, whose fast growth could plausibly have exposed more than estimated, classic herd immunity thresholds, served as a bounding, Epidemic Resistance Line (ERL) that predicted the speed of later resurgent epidemics following the relaxation of NPIs and during seasonally-driven epidemics in the fall. The NYC epidemic resistance line funneled growth rates to zero on a timescale of deaths, and in this way also served as a means of estimating the mortality burden at peak for thousands of US counties. This estimate was more accurate for counties whose states did not impose mask-wearing mandates, and was confirmed by inference of the vector field of all US counties’ trajectories, projecting the average US county’s trajectory from its point of maximum speed in the fall to a predicted mortality burden bounded above by the NYC line.

The corroboration of the NYC ERL and the real-time interpretation and visualizations of thousands of epidemic endpoints were done using a method we propose for graphical comparison of COVID-19 epidemics. In our method, disease burden, *D*, functions as a timescale for comparing of epidemics much like mileage serves as a timescale for comparing cars; the interpretation of burden as a timescale allows *r*(*D*) visualizations to be read from left to right, tagged by contemporary interventions, and aligned with other epidemics taking place at different calendar dates. These plots combine with the inferences of vector fields in (*r, D*) space to reveal mean-field patterns such as the funneling of growth rates to endpoints where *r*(*D*) = 0.

Our study has limitations. First and foremost, our visual method does not estimate the relative importance of immunity, behavioral changes, and other factors resulting in similar mortality burdens at asynchronous peaks. While hundreds of trajectories of less-mitigated epidemics followed a predictable pattern that lends itself naturally to hypotheses of herd immunity, alternative explanations exist. We use herd immunity thresholds to conceptually motivate the use of NYC as an ERL, but in our approach we remain agnostic about the relative importance of natural and waning immunity, behavioral changes, heterogeneity in susceptibility and transmission reducing attack rates at herd immunity, decreased infection fatality rates, and other causes of early endpoints that coincidentally aligned with the NYC predictions. To this point, NYC saw rising cases and the accumulation of deaths in the fall, which would reject a herd immunity hypothesis under a strict definition of herd immunity without waning of immunity and without novel variants with heightened transmissibility or immune evasion. More precise studies of the physiological, behavioral, virological, and seasonal drivers of COVID-19 transmission are needed to understand the mechanisms behind our effective bounds on transmission and death burden, as reported in this study.

Additionally, it is not possible to test the hypothesis that a region’s epidemic is “over” any more than a blindfolded passenger could test that their stopped cart is at the bottom of the world’s valley and not just a local valley. At the time of this writing, for example, the introduction of a variant of concern - B 1.1.7 - caused growth rates of cases across counties in Michigan that exceeded the NYC line. We would expect a more transmissible variant to exceed our estimated upper bounds on epidemic speed set by less transmissible variants, and we hypothesize this method applied to early B 1.1.7 epidemics may prove useful for estimating the epidemic potential and mortality burden for epidemics driven by variants of concern with increased transmissibility [30]. Waning immunity, the arrival of new susceptible individuals through birth and migration, the evolution of novel strains, and annual patterns in the seasonal forcing of transmission all require deliberate choice of date ranges over which we compare epidemics.

Our time range of interest was from the start of the COVID-19 pandemic in the US through the subsequent fall and winter epidemics, ending at the start of February 2021 after most US counties had peaked and prior to when more transmissible strains reached high prevalence and altered epidemic dynamics. This time range contains enough information for meaningful comparisons on the outcomes of epidemics from SARS-CoV-2 strains, but the sensitivity of our approach and its interpretations to the choice of timescale needs to be considered and, in each case, carefully justified. We encourage researchers to evaluate our approach with historical pandemics such as pandemic influenzas [31], seasonal influenza that arrive at natural local endpoints and are followed by resurgences every year [32], or the recent epidemics of Ebola [33] that were contained through management interventions. Applying the visualization comparisons and the associated analytical approaches we present here across more epidemics may improve the utility of this approach and help the scientific and management communities understand its uses and limitations for real-time comparison of epidemics of novel pathogens with pandemic potential.

An additional limitation is our use of mask-wearing mandates as a proxy for regulatory intensity where many regional, demographic, and other confounds exist, as they do in any ecological study [34]. Mask-wearing mandates at the state level may not correspond neatly to transmission-reduction at the individual level due to variation in statewide enforcement, county policies, business practices, and individual adherence to mask-wearing and physical distancing recommendations. We use per-capita deaths as a proportional measure of cumulative incidence, yet we don’t adjust for different demographics across regions or, more precisely, different demographics of the infected population. Differences in the correlation between the probability of infection and risk can change the infection fatality rate across regions and reduce the suitability of unadjusted deaths per-capita as a comparable measure of cumulative incidence. We also assume that infection fatality rates are similar across time. In reality, non-stationarity in the infection fatality rate from effective treatments or time-varying protection of vulnerable populations would require similar risk-adjustments to compare epidemics taking place at different points in time. Finally, while we feel that comparing the mortality burden at the final winter peak is a useful point of comparison in the context of understanding variation in *r*(*D*) trajectories, we note that these findings are not final estimates of mortality outcomes as later peaks, data-dumps, and other delayed effects can change the relative mortality burden across counties.

Our methods and analyses have many other limitations including regional and temporal biases or trends in case and death ascertainment, simplifying assumptions of similar infection fatality rates across regions & times used in our models motivating ERLs, and statistical methods used to estimate growth rates. Yet, despite these limitations, we are sensitive to the fact that public health and government officials have to act on multiple fronts, in real-time and in the face of uncertainty, to contain and control local outbreaks of pandemic pathogens to avoid overwhelming medical services and bridge the time until definitive pharmaceutical measures such as vaccines can be deployed, all while managing competing risks and considering the costs of their interventions. We present our analytical method and its visualizations of the US COVID-19 epidemics to harness the power of comparative epidemiology to give these decision makers insight into the potential speed and burden of epidemics in their locales that they may attain through relaxation of interventions to weigh against competing risks. Applying this approach to SARS-CoV-2 provides suggestive evidence that many US counties may have reached natural epidemic endpoints in the winter of 2020-2021 due to unknown combinations of behavioral changes, immunity, and waning seasonal forcing. Notably, we find these endpoints predicted well by our use of epidemic resistance lines occurred at COVID-19 mortality burdens significantly less than those estimated from the product of classical herd immunity thresholds and serosurvey-based estimates of infection fatality rates. The similar mortality burdens at the peak of confirmed cases may be due to a common level of mitigation across counties with vastly different population structures, NPIs, attitudes towards social distancing, and more. If later physiological studies find these endpoints of COVID epidemics are consistent with herd immunity, we hypothesize from equation 4 that it will be through an unknown mix of heterogeneity in transmission and susceptibility producing lower attack rates at herd immunity [15, 16, 17, 18] and/or lower infection fatality rates evident in the discovery of large populations of seronegative patients who have T-cells specifically recognizing SARS-CoV-2 [22, 23].

## Methods

### Data sources

We used the R package COVID19 [6] which compiled data from many sources. Through the R package COVID19, we primarily use the John’s Hopkins CSSE dataset [35] for cases at the country level, The COVID-19 Tracking Project for data at the US state level, and the New York Times github repo for US county-level data (https://github.com/nytimes/covid-19-data). All analyses are run in R version 4.0.2 and available at https://github.com/reptalex/COVID_ERL.

### Growth rate estimation

Growth rates were estimated from case data using a negative binomial state-space model with the R package *KFAS* [36]. Time-series outliers, detected by unusually large short-term jumps, were removed using the R package *tsoutliers* using an ARIMA(p,d,q) model fit via maximum likelihood where p=1, d=1, and q=2 were selected based on the Akaike information criterion [37].

We model the case counts for region *i* using a negative binomial generalized linear state space model. Specifically, for region *i*, we model the case counts at time *t* as:

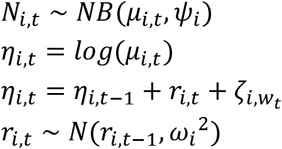

where *ψ*_*i*_ is the region’s negative binomial dispersion of case counts, *μ*_*i,t*_ the region’s time-varying negative binomial mean of case counts, *r*_*i,t*_ a unit-coefficient autoregressive term capturing unexplained drift in the average case counts over time, also interpretable as the exponential growth rates of cases, and 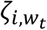 a day-of-the-week fixed effect whose magnitude depends on the region. The model was initialized with the prior *η*_*i*,0_ ∼ *N*(0,10) and *r*_*i*,0_ ∼ *N*(0,1).

### Choice of lag to align case growth rates and deaths per-capita

The negative binomial state space model provides a reliable instantaneous estimate of the exponential growth rate, *r*(*t*). However, to construct a timescale of cumulative incidence for comparison across regions, *r*(*t*) must be compared to lagged deaths, *D*(*t + ι*), for a lag, *ι*, equal to the average difference in time between when people exposed on a given day would be reported as a case versus when they would be reported as a death, or the difference from case reporting to death reporting for a cohort. Doing so ensures deaths per-capita are approximately proportional to cumulative incidence at the time of exposure for cases whose growth rate is being measured.

We found our estimated lag, *ι*, using the planning scenarios for COVID from the United States Center for Disease Control and Surveillance (CDC) as of September 9, 2020 [38]. The average lag from onset to seeking care was estimated by quantile matching a log-normal distribution to have 35% of the distribution below 2 days and 82% of the distribution less than or equal to 8 days, resulting in an average 5.3 day lag from onset to seeking care [39]. The lag from onset to death was estimated by log-normal quantile matching for the interquartile range in the CDC planning scenarios, and taking a weighted average of the resulting mean time from onset to death where weights were equal to the proportion of total deaths from that age group as of September 9, 2020. The resulting average lag from onset to death was 16 days. Based on the 5.3 day lag from symptom onset to case detection and a 16 day lag from symptom onset to death, we compare estimated growth rates *r*(*t*) to deaths per-capita *D*(*t + τ*_*D*_) for a *τ*_*D*_ = 11 day average difference from case to death reporting within a cohort exposed on the same day.

### Summer US state trajectories

Wikipedia timelines, news reports and, where possible, the executive orders, of interventions in place across six US states – Arizona, California, Florida, Georgia, Louisiana, and Texas – from the beginning of the US pandemic until the end of August, 2020 were collected and summarized in table S1: **“**Summer intervention timelines in focal US states”. Growth rates are estimated as mentioned above, deaths per-capita lagged by 11 days, and trajectories from March 1 until August 31 are plotted. Inset case and growth rate trajectories over time are also plotted to include the familiar timescale of calendar dates.

### Peak and prominence estimation

Peaks were identified as dates when *r*(*t*) estimates switched sign from positive to negative. The prominence of a peak was estimated as the ratio of the model’s expected case counts at the peak to the expected case counts at the preceding valley. Plots in figure 4 illustrating many peaks of the top-5 most populous US counties outside NYC show all identified peaks regardless of prominence for completeness, and plots showing the many peaks of US counties over time are limited to peaks with 3x the prominence of preceding valley.

### Mortality outcomes based on statewide mask-wearing mandates

Based on reporting [29, 40], the following states were defined as not having state-wide mask-wearing mandates: Arizona, Florida, Georgia, Idaho, Missouri, Nebraska, Oklahoma, South Carolina, South Dakota, and Tennessee.

To estimate the mortality burden at the final peak, we first determined the latest peak prior to 2021-02-01 with a prominence greater than 3x the case counts of the preceding valley. This resulted in 2,295 counties for which we had *D*^*\**^ estimates for the final peak. Using a binomial generalized additive model, we predicted the log-odds of a member of the population dying due to COVID-19 with a piecewise cubic spline of log population size and whether/not the county was found in a state with a state-wide mask-wearing mandate.

To predict the counter-factual mortality burden, the model estimated above was used to predict the mortality of every county assuming the converse policy was in place in its state. The difference between counter-factual deaths and observed deaths was summed across all counties based on state-wide mask-wearing policy, excluding New York state as 80% of the COVID deaths in NY occurred in the first wave prior to the likely efficacy of mask-wearing, resulting in our estimated 29,000 lives saved across mask-wearing states and 7,700 excess deaths across states without mask-wearing mandates.

### Vector field inference and integration

Implicitly defining logistic growth by *dN*/*dt* defines a vector field of population changes, Δ*N*, over time increments, Δ*t*. As a timescale, per-capita deaths define a vector field of expected transmission changes, Δ*r*(*t*), over incremental changes in per-capita deaths, Δ*D*, including log-scaled Δlog (*D*). To estimate the vector field of all US counties’ *r*(*D*) trajectories, we first computed the changes in growth rate estimates from day to day, Δ*r*_*t*_ = *r*(*t +* 1) − *r*(*t*) and the associated change in the logarithm of deaths per-capita from day-to-day, Δ log(*D*_*t*_) = log(*D*(*t + τ +* 1)) − log (*D*(*t + τ*)). These allowed empirical estimates of the instantaneous slope of every counties’ *r*(*D*) trajectories through Δ*r*/Δ log(*D*). Two separate generalized additive models were ran predicting Δ*r* and Δlog (*D*) as

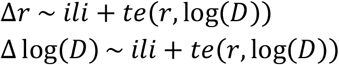

Where *s*() denotes a polynomial cubic spline and *te*() a tensor spline and *iιi* is the historical average proportion of patients visiting outpatient providers with influenza-like illness across states for the corresponding week of the year. These models predict the growth rate differentials, 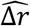, and log per-capita death differentials, 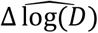, at every point in (*r*, log(*D*)) space. A grid of n=100 points equally spaced across *r* ∈ [−0.2,0.6] and log(*D*) ∈ [log(10^−5^), log(0.004)] was used to compute the epidemic trajectory vectors 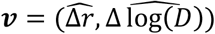. These vectors were converted to unit vectors and sized to a common length. These models are not unlike the state-space models used to estimate the changes of cases over time in the negative binomial state-space models above. The equation to infer growth rates, for example, can be re-written as

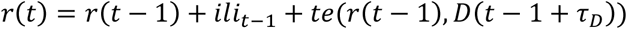

which defines a nonlinear and non-stationary state-space model over the timescale of deaths.

Integration of predicted trajectories was done by initializing *D*(0) = 10^−5^, *iιi* = 3.8% (the historical maximum, which occurs on week 52), and sampling 100 equally spaced values of *r*(0) ∈ [0,1]. Euler integration sequentially used the estimated differentials to compute trajectories until *D* = 2 × 10^−3^ or *r* = −0.1.

To simulate a US fall maximum trajectory, the maximum growth rate after August 1 for every county was identified and its corresponding *D*(*t +* 11) extracted. The median of these maximum growth rates and the median of their corresponding deaths per-capita defined a point in (*r, D*) space; the numerically integrated trajectory closest to this point is highlighted and labelled “US fall max”.

## Data Availability

Data are available online through the R package COVID19, and we make our scripts available at the github repo: reptalex/COVID_ERL

https://github.com/reptalex/COVID_ERL

## Supplemental Information

### 1. Analysis of epidemics and their endpoints on the timescale of deaths

In this section, we analyze two common epidemiological models to motivate our plots of epidemic growth rates as a function of lagged deaths per-capita. The main value of these analytical approximations of *r*(*D*) trajectories is to point out that while herd immunity is defined by a single point – an attack rate where prevalence no longer increases - epidemic resistance lines define an entire surface bounding the speed of epidemics from the epidemic’s maximum speed until a population fatality rate at the herd immunity threshold (or effective herd immunity threshold). We also show in a simulation how our approximations for ERLs in these model systems work reasonably well and, more importantly, how the empirical epidemic resistance lines from simulations of unmitigated epidemics outperform our approximations, thereby motivating the value of empirical epidemic resistance lines in real-world epidemics as potentially superior to analytical approximations from simple models (much as approximations of herd immunity thresholds are oversimplifications, so too are approximations of ERLs).

### SIR model

For an SIR model for which births and non-infected mortality is negligible over the course of an epidemic and *S + I + R* = *N* for all time, written as a system of differential equations:

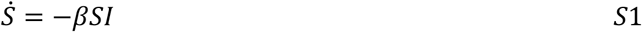

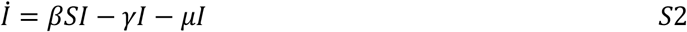

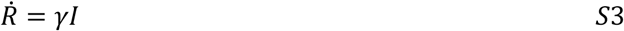

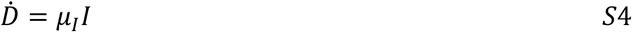

we note that if cases are confirmed proportional to prevalence then the expected growth rate of cases will be the growth rate of prevalence,

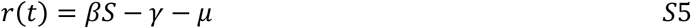

and by defining cumulative incidence, *Z* = *N* − *S*, we can write growth rates as a function of cumulative incidence

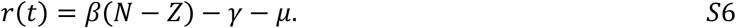

Expressing growth rates as a function of deaths can be done by approximating cumulative incidence as a function of deaths. Defining the instantaneous incidence *z*(*t*) = *βSI*, we can write

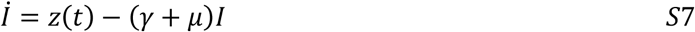

which is a linear first order differential equation with solution

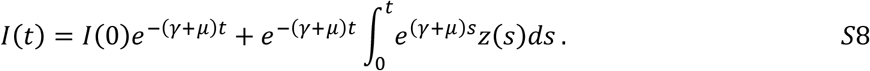

Setting *τ* = *t* − *s*, assuming *I*(0)*e*^−(*γ+μ*)*t*^ ≈ 0, and multiplying and dividing the integrand by *γ + μ* yields

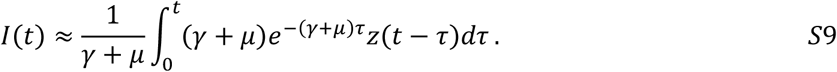

The right-hand side is proportional to an exponentially weighted moving average of *z*(*t*). Alternatively, this can be seen as the expectation of *z*(*t* − *τ*) for a random variable *τ*∼exp (*γ + μ*). The latter can be approximated by substituting the expected value of *τ* into *z*(*t* − *τ*), yielding

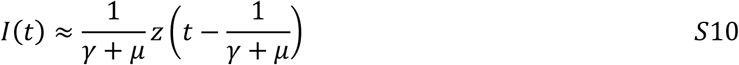

or that the prevalence is approximately proportional to a lagged incidence curve. Combined with the differential equation for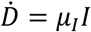 and integrating to the present time produces a relationship between the cumulative deaths and cumulative incidence

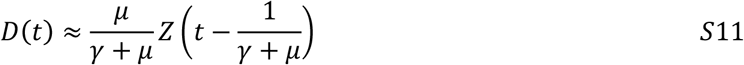

where deaths today approximate cumulative incidence in the past. Taking the same approach for *R*(*t*) results in the approximation that 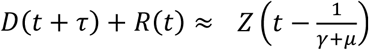, i.e. that everyone infected (*γ + μ*)^−1^ days ago has either died or recovered. The time shift for which deaths approximate cumulative incidence is the expected time it takes for an infected individual to either recover or die.

Inserting our approximation of cumulative incidence as a function of lagged deaths (equation S11) into our equation for the growth rate in cases as a function of cumulative incidence (equation S6) yields

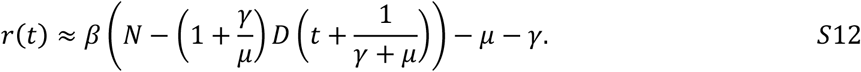

Defining *D*_*ι*_ as the appropriately lagged deaths in the preceding equation one can rearrange terms with a little algebra to re-write *r*(*D*_*ι*_) in a useful slope-intercept form:

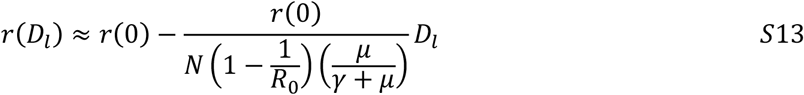

where *r*(0) = *βN* − *μ* − *γ* is the initial growth rate of the epidemic with one infected individual and 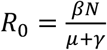 for an SIR model. Equation S13 can be simplified even further by recognizing 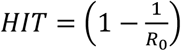 is the herd immunity threshold and 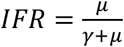 is the infection fatality rate, resulting in

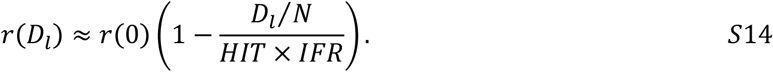

The equation above says that when deaths per-capita today imply that (*γ + μ*)^−1^ days ago cumulative incidence was equal to the herd immunity threshold times the infection fatality rate, we will have expected prevalence (*γ + μ*)^−1^ days ago to peak.

Our findings from an SIR model have little immediate use for analyzing COVID epidemic trajectories across countries given nonzero incubation periods and a variety of other assumptions that don’t hold in real populations, but the final equation provides clear intuition about epidemic resistances line and the timescale of deaths. The y-intercept defines the initial growth rate of COVID prevalence in that region and the x-intercept reveals the herd immunity threshold. Increasing the infection fatality rate, all else being equal, will reduce the rate of descent of *r*(*D*_*ι*_) by increasing the rate at which deaths accumulate. The peak of the epidemic where *r*(*D*_*ι*_) = 0 is determined by the product of herd immunity threshold and the infection fatality rate.

Reducing transmission by interventions will reduce *r*(*D*_*ι*_) relative to the unmitigated scenario, resulting in a departure from a linear trajectory – this can also be easily seen by taking the time derivative of equation S6 to yield

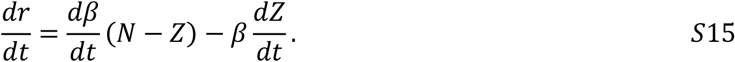

Or, using *D*_*ι*_(*t*) as our definition of a timescale, yields

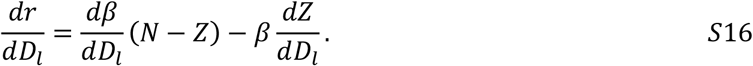

The linear approximation for *r*(*D*) is from the constancy of 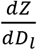 in our approximation in equation S11. However, under interventions that successfully reduce transmission, *β* will not be constant. having non-stationary transmission yielding non-constant 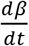 will introduce nonlinearities both through 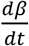 and the way 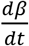 is modulated by its product with *N* − *Z* (i.e. increasing cumulative incidence will decrease the effect of 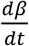 because the varying rate of transmission is dampened by the reduced population of susceptibles).

The density-dependent SIR model above is used to provide intuition on the mechanism by which *r*(*D*) trajectories define empirically observable effective herd immunity thresholds along an entire surface, rather than a final attack rate alone. Epidemic resistance lines may be expected from natural epidemics assuming transmission rates decrease with increasing cumulative incidence and real-time estimates of burden can be made approximately proportional to cumulative incidence.

More complicated epidemiological processes will have different functions, *f*_*I*_ for transmission, including with nonlinear functional forms and more complex dependencies on multiple compartments. Wallinga & Lipsitch [9] illustrated that while different models have different relationships between exponential growth rates *r*(*t*) and effective reproductive numbers *R*_*t*_, these models are unified by a common, Lotka-Euler equation connecting the two: the effective reproductive number is the Laplace transform of the serial interval distribution, *g*(*s*) evaluated at the exponential growth rate:

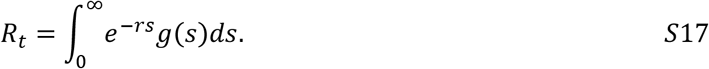

Rather than exhaustively explore ERLs of different model structures, we focus on the intuitive connection between exponential growth rates and transmission, between transmission and cumulative incidence, and between cumulative incidence and a measure of burden.

For illustrative purposes, we present two scenarios in which an early, less-mitigated epidemic’s trajectory would NOT define an ERL. In one scenario, if a less mitigated epidemic is in a population whose contact structure results in a lower baseline rate of transmission, then later epidemics with contact structures resulting in a higher baseline rate of transmission can exceed the early, less-mitigated empirically hypothesized ERL. Another scenario would be if an early, less-mitigated epidemic occurred at a time when seasonal forcing dampened transmission and later epidemics occurred when seasonal forcing amplified transmission. These scenarios share a common theme of heterogeneity in transmission of space or time, and early epidemics occurring at a place or time with low transmission which consequently does not bound transmission at other places or times. For COVID, we’ve either focused on the NYC epidemic that occurred at a similar high-point of seasonal forcing as later fall epidemics, or Sweden when compared to US summer epidemics at a similar, low point of seasonal forcing.

New York City’s densely populated built environment, high rates of public transportation use and more make it plausibly a worst-case-scenario for respiratory viral transmission, and so its use as an upper bound for other US counties may satisfy the requisite assumptions for careful use of ERLs to hypothesize maximum resurgent epidemic speeds and later epidemic endpoints.

**2. Table S1:**
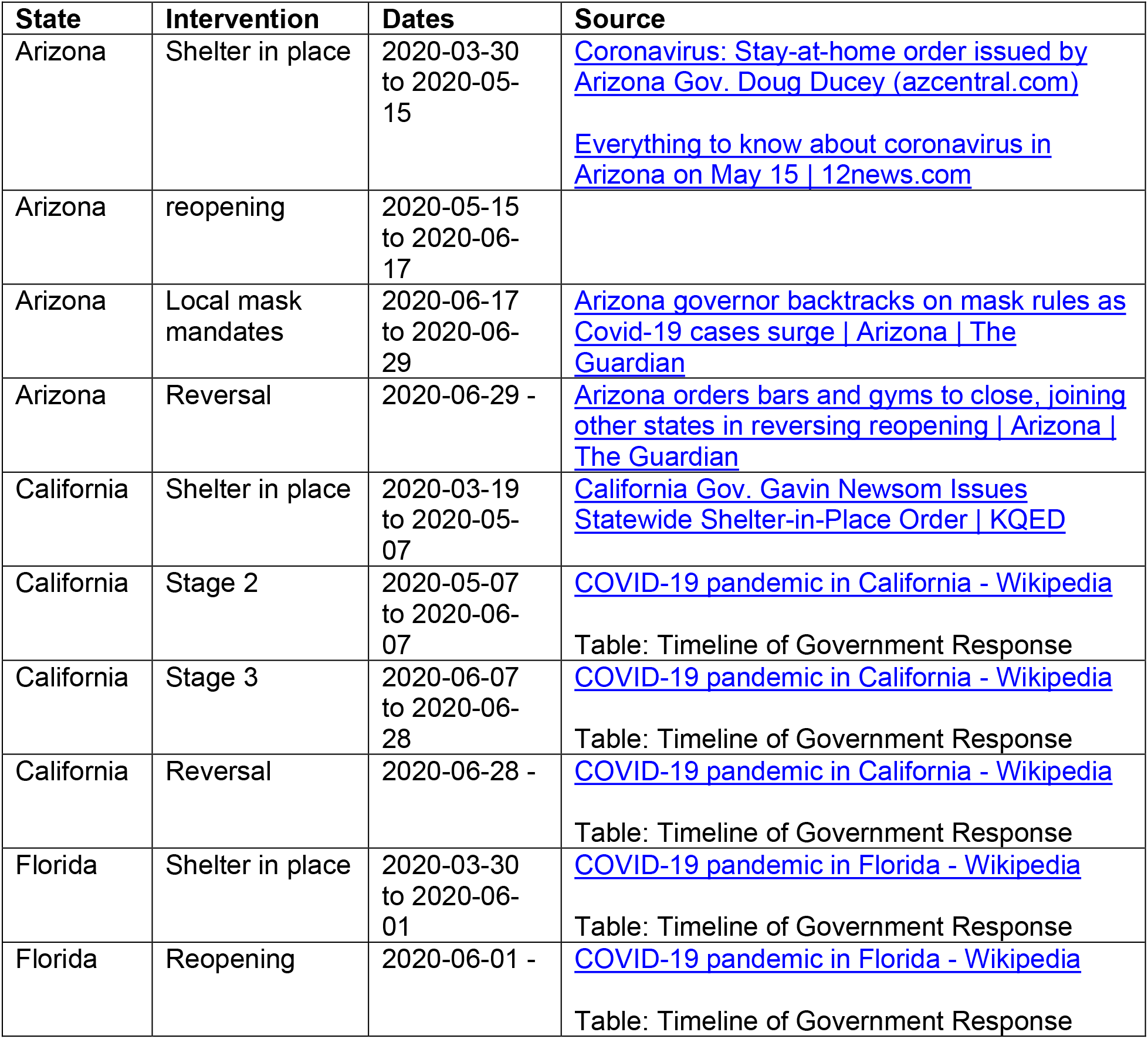
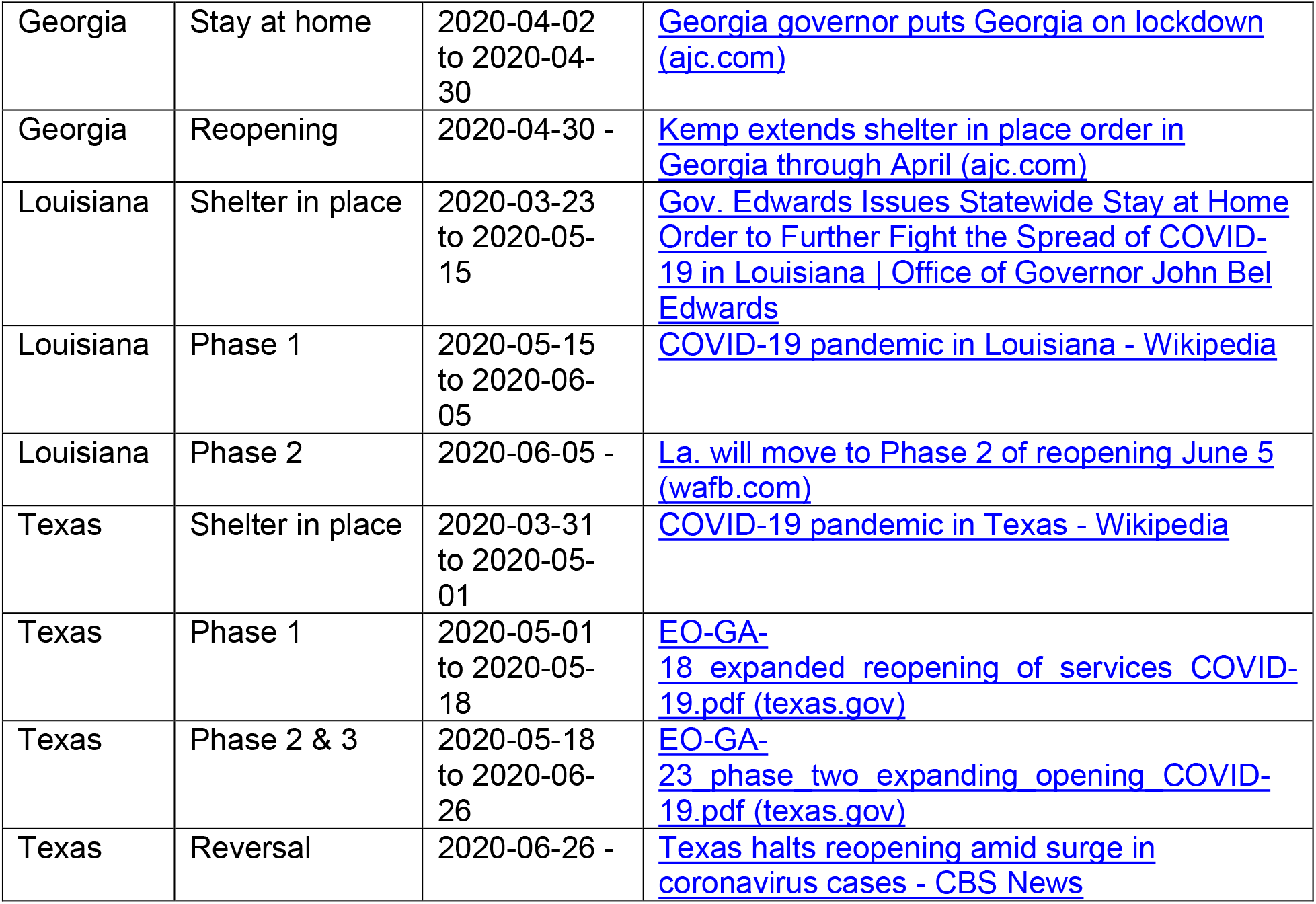
Summer intervention timelines in focal US states.

| State | Intervention | Dates | Source |
| --- | --- | --- | --- |
| Arizona | Shelter in place | 2020-03-30 to 2020-05-15 | Coronavirus: Stay-at-home order issued by Arizona Gov. Doug Ducey (azcentral.com)<br>Everything to know about coronavirus in Arizona on May 15 \| 12news.com |
| Arizona | reopening | 2020-05-15 to 2020-06-17 |  |
| Arizona | Local mask mandates | 2020-06-17 to 2020-06-29 | Arizona governor backtracks on mask rules as Covid-19 cases surge \| Arizona \| The Guardian |
| Arizona | Reversal | 2020-06-29 - | Arizona orders bars and gyms to close, joining other states in reversing reopening \| Arizona \| The Guardian |
| California | Shelter in place | 2020-03-19 to 2020-05-07 | California Gov. Gavin Newsom Issues Statewide Shelter-in-Place Order \| KQED |
| California | Stage 2 | 2020-05-07 to 2020-06-07 | COVID-19 pandemic in California - Wikipedia<br>Table: Timeline of Government Response |
| California | Stage 3 | 2020-06-07 to 2020-06-28 | COVID-19 pandemic in California - Wikipedia<br>Table: Timeline of Government Response |
| California | Reversal | 2020-06-28 - | COVID-19 pandemic in California - Wikipedia<br>Table: Timeline of Government Response |
| Florida | Shelter in place | 2020-03-30 to 2020-06-01 | COVID-19 pandemic in Florida - Wikipedia<br>Table: Timeline of Government Response |
| Florida | Reopening | 2020-06-01 - | COVID-19 pandemic in Florida - Wikipedia<br>Table: Timeline of Government Response |
| PRELIMINARY - NOT PEER REVIEWED |  |  |  |
| Georgia | Stay at home | 2020-04-02 to 2020-04-30 | Georgia governor puts Georgia on lockdown (ajc.com) |
| Georgia | Reopening | 2020-04-30 - | Kemp extends shelter in place order in Georgia through April (ajc.com) |
| Louisiana | Shelter in place | 2020-03-23 to 2020-05-15 | Gov. Edwards Issues Statewide Stay at Home Order to Further Fight the Spread of COVID-19 in Louisiana \| Office of Governor John Bel Edwards |
| Louisiana | Phase 1 | 2020-05-15 to 2020-06-05 | COVID-19 pandemic in Louisiana - Wikipedia |
| Louisiana | Phase 2 | 2020-06-05 - | La. will move to Phase 2 of reopening June 5 (wafb.com) |
| Texas | Shelter in place | 2020-03-31 to 2020-05-01 | COVID-19 pandemic in Texas - Wikipedia |
| Texas | Phase 1 | 2020-05-01 to 2020-05-18 | EO-GA-18 expanded reopening of services COVID-19.pdf (texas.gov) |
| Texas | Phase 2 & 3 | 2020-05-18 to 2020-06-26 | EO-GA-23 phase two expanding opening COVID-19.pdf (texas.gov) |
| Texas | Reversal | 2020-06-26 - | Texas halts reopening amid surge in coronavirus cases - CBS News |

### 3. Seasonal Forcing

**Figure S1.**
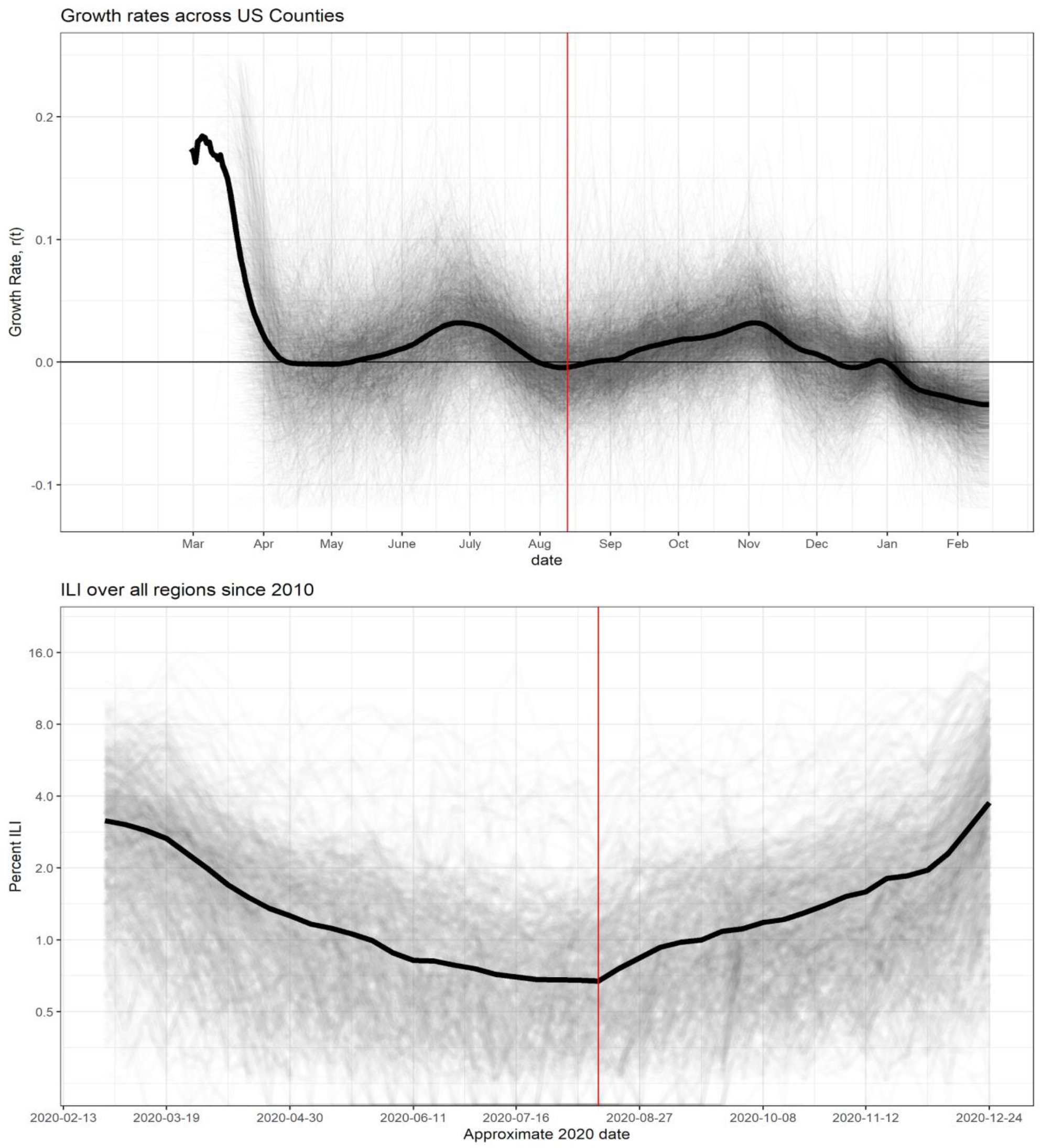
The first series of local endpoints were peaks in July-August. Starting roughly week 33, the growth rates of cases across us counties showed a synchronized rise. This transition to a fall epidemic began at roughly the 2020 equivalent of when historical average ILI visits to outpatient providers rose, suggesting this rise is consistent with seasonal forcing known to underlie the dynamics and seasonal epidemics of other respiratory pathogens causing influenza-like illness.

